# Long COVID and Work in the UK: Challenges, Support and Perspectives

**DOI:** 10.1101/2025.05.08.25327220

**Authors:** Hua Wei, Sarah Daniels, Ruth Wiggans, Anna Coleman, Donna Bramwell, Damien McElvenny, Davine Forde, Martie van Tongeren

## Abstract

**Aim:** Long COVID (LC) presents significant challenges for working age individuals, leading to major inequalities in access to work, employment and relevant support. This study investigates the workplace support provided to people with Long COVID (PwLC) in the UK, focusing on their return- to-work (RTW) experiences. It encompasses perspectives from both PwLC and managers of PwLC.

**Methods:** Semi-structured interviews were conducted with 20 PwLC and two managers experienced in managing employees with LC. Thematic analysis was performed using NVivo14.

**Findings:** This qualitative research explored barriers and facilitators to supporting PwLC’s RTW. LC is characterised by a wide range of mostly “invisible” and fluctuating symptoms and unpredictable recovery trajectories during which relapses can occur. Existing support mechanisms for RTW with LC include phased return, reduced hours, Occupational Health services, work adjustments, and government support. However, the study identified challenges in implementing these measures, such as unrealistic phased return plans, managers neglecting advice or guidance (e.g. from Occupational Health), unsuitable work adjustments and the burden of navigating government support. The financial impact of reduced hours or sick leave was one of the main reasons for returning to work. Both PwLC and managers highlighted significant gaps in knowledge, training, policy and guidance for RTW support, emphasising the need for tailored support. Managers reported limited resources and inflexible policies as main challenges, which they addressed through creative solutions.

**Conclusion:** This qualitative study highlights potential barriers, challenges and gaps in supporting PwLC’s RTW. To ensure equitable access to work for PwLC, a flexible and personalised approach is crucial, given the variability in LC symptoms and recovery rates. RTW support that fails to accommodate these characteristics may exacerbate symptoms or cause relapses. A supportive work environment is essential, as LC symptoms can be invisible and concerns about stigma may prevent PwLC from communicating openly and seeking support. Lack of resources is a major barrier for managers in supporting PwLC. Effective government support can potentially fill this gap but must be well-designed and implemented to reduce the burden on applicants.

## Introduction

A significant number of individuals continue to suffer from persistent symptoms after initial infection with COVID-19, a condition known as Long COVID (LC) or Post-Acute COVID-19 Syndrome (PACS) [1, 2]. LC is characterized by a wide range of symptoms, including fatigue, cognitive dysfunction, respiratory issues, musculoskeletal pain and other symptoms, lasting longer than 12 weeks [3, 4]. As of March 2024, approximately two million people were experiencing LC (3.3% of the population) [5] and a recent study found that almost 5% of general practice (GP) patients were estimated to have LC [6]. Many of these individuals are of working age and continue to experience debilitating symptoms [7].

Studies have shown that LC can severely impair work functioning, leading to reduced productivity, increased absenteeism or presenteeism, and, in some cases, job loss or early retirement. For instance, a survey by the Trade Union Congress found that a large proportion of workers with LC reported fatigue, brain fog, and other symptoms that hindered their ability to perform their job duties effectively [8]. The fluctuating and unpredictable nature of LC symptoms further complicates the RTW process, making it challenging for both employees and employers to manage [9, 10].

There are examples of effective support and accommodation that can facilitate the RTW process for individuals with LC [11, 12]. Flexible work arrangements, such as remote work, regular breaks and reduced hours, have been identified as crucial in helping workers manage their symptoms while maintaining employment [13, 14]. Supportive management practices, including regular check-ins, empathy, and understanding of the condition, can significantly improve the work experience for those with LC [10]. Additionally, organisational policies that recognise the unique needs of LC sufferers and provide tailored accommodations can facilitate RTW for PwLC [9, 15, 16].

Nevertheless, research on the experiences of workers with LC showed unsatisfactory experiences and continuous challenges. These include limited adjustments to accommodate physical and cognitive impairment, unmet needs for flexible work arrangements, and psychological impacts in the long term [2, 8]. Workers often struggled with the stigma associated with LC, feeling misunderstood or unsupported by colleagues and management [9, 17, 18]. The lack of awareness and understanding of LC among employers can lead to inadequate workplace accommodations and support, exacerbating the difficulties faced by affected individuals [1, 19].

This qualitative study aimed to explore the types of support that can help workers with LC to maintain employment, as well as the specific challenges they encounter. By drawing on perspectives from both PwLC and managers experienced in supporting them, we were able to facilitate discussions with both groups to identify barriers to effective support. Our objective was to provide practical recommendations for employees, employers, and policymakers, to promote more equitable and supportive workplaces for individuals navigating the impacts of LC.

## Methods

We carried out semi-structured interviews (February to June 2023) with 20 PwLC and two managers, all based in the UK, who had experience in managing employees with LC. We followed the Consolidated Criteria for Reporting Qualitative Studies (COREQ) to report the findings [20]. The checklist can be found in Supplementary File 2. Ethical approval was obtained from the University of Manchester Research Ethics Committee (Ref: 2024-17658-32518).

### Recruitment and Data Collection

PwLC were defined either as: 1) currently or previously having had LC, and 2) being employed at the time of index COVID-19 infection. For managers, the inclusion criteria required current or recent experience in managing employees with LC in a workplace setting. PwLC were recruited through patient support groups for LC and the project was advertised on social media. Managers were recruited through the Greater Manchester Chamber of Commerce, regional networking events for employers, and local healthcare organisations. PwLC was the main focus of recruitment and we stopped recruiting when the research team agreed that data saturation was reached. Materials for recruitment, study information and interview schedules were agreed within the study team which included our patient and public involvement (PPI) representative. Potential participants were sent an Expression of Interest (EOI) form on Qualtrics. Participants were then approached by researchers (HW/SD) via email and sent a Participant Information Sheet (PIS) at least one working day before the interview. Written consent was obtained either through returning a signed consent form or through verbal consent recorded immediately before the interview.

Two trained researchers (HW and SD) experienced in qualitative research conducted all the interviews. Participants were asked to complete a questionnaire which asked for basic demographics (sex, age, ethnicity, marital status, and educational attainment), occupational history, self-reported level of disability, and time off work related to LC diagnosis. Interviews then followed a question schedule informed by a literature review and developed in collaboration with the project PPI representative. Interview questions for PwLC asked about their experience with LC including specific LC symptoms, work history, post-LC employment and relevant changes, support received and suggestions for improvement. Interview questions for managers or employers included information about the organisation, policies and practice in relation to supporting LC workers, their own experience in supporting PwLC, barriers and facilitators of RTW and suggestions for improvement. Open-ended questions were included to allow the participants to freely discuss topics they considered important and relevant to the research [21]. The interview schedules can be found in Supplementary File 1. The interviews lasted 30-60 minutes, were audio recorded and conducted either online via Zoom or Microsoft Teams teleconferencing platforms, or in-person.

### Data analysis

Interview audio recordings were anonymised and transcribed by a University-approved transcription service.

Thematic analysis was performed using NVivo14 software. An initial codebook was developed after two coders (HW and SD) independently coded five transcripts. Inter-coder reliability was assessed, yielding a high agreement rate of 99% and an average kappa coefficient of 0.62, which falls within the “good” range for strength of agreement (0.61–0.80) [22]. Coding was done inductively following the latent approach [23, 24] identifying and examining words, phrases and sentences, and assigning new codes that reflected their meaning. Related codes were grouped into sub-themes and then organised into overarching themes to form the initial codebook. This codebook was reviewed and discussed with the wider research team leading to an iterative re-coding process. This continued until the team reached a consensus on the final coding structure. Following this, the researchers independently coded the remaining transcripts using the coding structure.

## Results

### Demographics

Between February and June 2024, we interviewed 20 PwLC and two managers experienced in managing PwLC. The table below shows the demographics of the PwLC interviewees (Table 1). The majority of participants were women (n=15, 75%) aged over 40. All but one worked in professional or technical roles with high levels of educational attainment. While most worked in medium or large companies, three participants worked in small organisations with fewer than 10 employees, and two were self-employed. All but one participant was White British. Participants reported a mean time off work due to LC of 11.3 (range 0 – 29) months. Most (n=15, 75%) participants were working at the time of study recruitment. Twelve participants were based in Northwest England, with the rest from other parts of England or Scotland.

**Table 1:** Demographics of interviewed PwLC.

| Particip | Gender | Age | Company | Work | Ethnicity | When LC | Time off | UK |
| --- | --- | --- | --- | --- | --- | --- | --- | --- |

| ant number |  | Range | size<sup>1</sup> | ing<sup>2</sup> |  | develope d | work, months | Region<sup>3</sup> |
| --- | --- | --- | --- | --- | --- | --- | --- | --- |
| 1 | M | 60-69 | Large | Y | British White | Jul-20 | 7 | NW |
| 2 | F | 50-59 | Large | Y | British White | NR | 4 | NW |
| 3 | F | 50-59 | Small | Y | British White | Oct-21 | 29 | NW |
| 4 | F | 50-59 | Large | Y | British White | Aug-21 | 21 | NW |
| 5 | F | 30-39 | Large | Y | British White | Nov-23 | 0.5 | NW |
| 6 | M | 50-59 | Large | Y | British White | Jan-22 | 0 | NW |
| 7 | F | 40-49 | Medium | N | British White | Aug-22 | 18 | NW |
| 8 | F | 40-49 | Large | Y | British White | Feb-22 | 10 | YH |
| 9 | M | 30-39 | Self employed | N | Ashkenazi | Dec-21 | 27 | EM |
| 10 | F | 40-49 | Small | Y | British White | Nov-21 | 0.3 | NW |
| 11 | F | 40-49 | Small | N | British White | Mar-22 | 10 | NW |
| 12 | F | 30-39 | Large | Y | British White | Dec-23 | 3.5 | NW |
| 13 | F | 50-59 | Large | Y | British White | May-21 | 10.5 | EM |
| 14 | M | 40-49 | Large | Y | British White | Sep-23 | 2 | YH |
| 15 | F | 50-59 | Large | Y | British White | May-22 | 24.6 | EM |
| 16 | F | 20-29 | Large | Y | British White | Mar-20 | 18 | GL |
| 17 | F | 50-59 | Large | N | British White | Mar-21 | 11 | EM |
| 18 | F | 50-59 | Large | Y | British White | Dec-21 | 27 | Scotland |
| 19 | M | 30-39 | Self employed | N | British White | Mar-22 | NR | NW |
| 20 | F | 40-49 | Medium | Y | British White | Oct-21 | 3.5 | NW |

<sup>1</sup>Large businesses – 100+ employees, Medium businesses – 10 - 99 employees, Small businesses – < 10 employees. <sup>2</sup>Working (yes or no) at time of study recruitment. <sup>3</sup>NW: Northwest, England; YH: Yorkshire and the Humber, England; EM: East Midlands, England; GL: Greater London, England. Abbreviations: NR – not reported

For the two managers, one worked for a large organisation in the capacity of occupational health and the other worked in a school as a head teacher.

### PwLC’s Perspective

#### LC symptoms and impact on work

The participants reported a broad range of symptoms and characteristics of LC, such as brain fog, fatigue, breathing difficulties, pain, post-exertional malaise, sleeping disturbances, speech impairments, gastroenterological and digestive problems. Most participants reported the fluctuation or relapse of LC symptoms as particularly challenging to manage and less understood by others (P2-5, P7, P10-11, P13-16, P19-20).

Living with LC had significant impacts on work. Participants reported reduced productivity when at work, working reduced hours, increased time off work, difficulties with commuting, and concerns about workplace safety, including assault at work (P13), feeling unsafe to travel as required by the job (P2), and concerns about patient safety where the participant was a medical practitioner (P15). Impacts on employment included non-renewal of fixed-term contracts due to LC (P11, P19), redundancy (P7), involuntary resignation (P13), voluntary resignation (P16), dismissal on ill-health grounds (P15), ill-health retirement (P17), and changes in contract conditions, e.g. from full-time to part-time (P3-4). Some participants described LC related presenteeism (P1, P6-7, P11, P13, P15-16, P18). The majority of participants reported being unfit or less fit for work. Some were advised by professionals, including healthcare providers, LC clinics, support groups and occupational health services, or colleagues (P4, P11-13, 15, 20), while others based their assessments on personal judgment (P1-2, P4-5, P7, P10, P12, P16, P19-20). Several participants described how they struggled with commuting either by driving or taking public transport (P1, P3, P8, P13-14), which added extra challenges to returning to in-person work.

#### Personal reasons for returning (or not) to work

The reasons for returning to work are categorised into personal and work-related factors. Personal reasons, such as financial pressures, are discussed first in this section. Work-related factors will be discussed later under the theme of “Workplace challenges for PwLC”.

Financial pressure was the most quoted personal reason for returning to work (P2, P10-12, P18, P20). P10 referred to the rising living costs:

> *“So, with the cost-of-living crisis and, obviously, my mortgage has gone up by a hundred and forty pounds a month, my gas and electric have gone up. So, I just couldn’t afford…[taking time off sick]”.*

For example, P18 said that:

> *“I am the main earner in the household, so if I lose my job there is quite a significant financial impact and we will have to sell our home and stuff like that”.*

Personal fulfilment, professional identity and the benefits to wellbeing were other important reasons for returning (P1, P4, P8, P11, P14, P16). For example, P8 said:

> *“I think for me it’s been a very big thing, it was, …my psychological wellbeing as well, because actually work is a big part of who we are, and I obviously trained for a long time, and I’ve done lots of exams, and enjoy my work and enjoy the contact with patients, and that’s a big part of me.”*

Some also considered the social benefit of work (P8, P14, P16) and job security (P3, P7).

Reasons for not returning to work were commonly associated with concerns about the impact of work or the workplace on LC symptoms and also on participants’ mental health. Some participants were just too poorly to go back to work or the original job role (P13, P15, P19), in one person this was attributed to repeated COVID-19 infection (P13). Some made the decision of not returning voluntarily because they were concerned that work would have exacerbated the symptoms or impeded their recovery (P7, P9, P15, P19). Some were willing to go back to work, but felt unable to do so due to concerns about managing the workload without the necessary adjustments, or worried they might be judged or misunderstood by their colleagues (P7, P13). Reflecting on her own experience and that of other PwLC, P7 summarised it well:

> *“… Because it [work] puts a lot of strain on you that you don’t know is there, like, knowing I have to be better by a certain point, and am I well enough to perform these actions, if I go back to work on phased return now is it going to make me more ill?*

> *…Having spoken to people with long COVID who have gone back to work, they seem to fall into two categories: they either just get more ill and are off work again, and aren’t even back to zero, are back to minus; or they have no life, they work, they go home, they sleep…*

> *And they struggle so much with colleagues judging, not understanding because we look fine, et cetera, et cetera. So I think not having that, for the present, I think is good for my health, it’s good for my physical health and my mental health.”*

#### Workplace support for PwLC

According to interviewed PwLC, support provided by workplaces primarily focussed on two themes: communication, recognition and understanding with and for PwLC; and work adjustments accommodating LC.

### Adjustments

Participants described workplace adjustments that helped them to return to or stay in work (Table 2), including autonomy, flexibility, or breaks, phased returns, reduced working hours, remote working or working from home (WFH), following advice or guidance (e.g. from occupational health, NHS or the Society of Occupational Medicine), and other workplace adjustments or arrangements, which included allowing time for appointments or attending support groups (P13), developing a personalised approach to align with the individual’s work capacity (P14), conducting regular check-ins to adjust workloads as needed (P16), arranging to share workloads (P18, P20), reducing physically demanding duties (P2, P3), changing job roles (P4, P5) and providing improved IT equipment (P8).

**Table 2:**
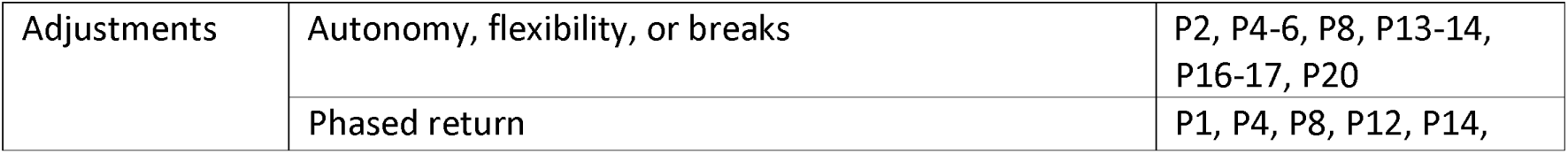
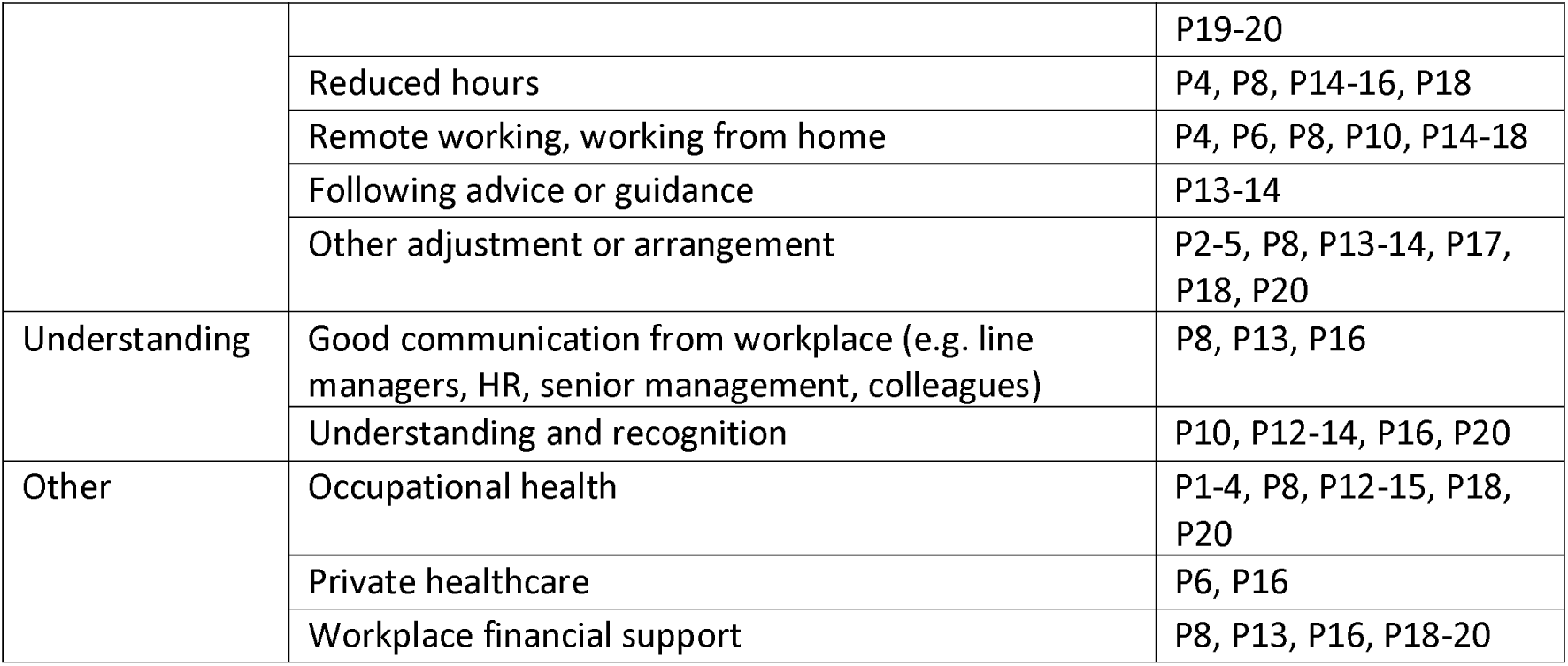
Workplace support for PwLC.

|  |  |  |
| --- | --- | --- |
| Adjustments | Autonomy, flexibility, or breaks | P2, P4-6, P8, P13-14, P16-17, P20 |
|  | Phased return | P1, P4, P8, P12, P14, |
|  |  | P19-20 |
|  | Reduced hours | P4, P8, P14-16, P18 |
|  | Remote working, working from home | P4, P6, P8, P10, P14-18 |
|  | Following advice or guidance | P13-14 |
|  | Other adjustment or arrangement | P2-5, P8, P13-14, P17, P18, P20 |
| Understanding | Good communication from workplace (e.g. line managers, HR, senior management, colleagues) | P8, P13, P16 |
|  | Understanding and recognition | P10, P12-14, P16, P20 |
| Other | Occupational health | P1-4, P8, P12-15, P18, P20 |
|  | Private healthcare | P6, P16 |
|  | Workplace financial support | P8, P13, P16, P18-20 |

### Understanding, Recognition and Good Communication

When managers, colleagues, and senior management showed that they really cared (P12), believed and understood what the participants were experiencing (P10, P12-13, P16), were willing to accommodate and make necessary allowances (P14), avoided adding pressure (P13), and maintained regular communication (P8, P13, P16), the participants felt supported by their workplace. For example, P20 reported:

> *“They were all brilliant when I returned to work. There were so many times when I would just say I’m so sorry that you’re having to do this, I’m so sorry, and they were like it’s absolutely fine, we’re just happy to have you back. So they were really willing to do whatever they could to help me with it.”*

P13 noted:

> *“I have a meeting with the manager, the business manager, once a month, who’s very supportive with the long COVID side of things and if there’s anything I need, she’s very good with that.”*

### Other Support

Additional support such as occupational health, private healthcare, and workplace financial support were also considered helpful. Among these, occupational health emerged as the most frequently discussed and consistently supportive mechanism in helping PwLC to remain in, or return to, work.

#### Workplace challenges for PwLC

PwLC encountered a range of challenges at different stages of their RTW journey. The sub-themes within this main theme included challenges when returning or preparing to return, challenges after returning, and environmental or organisational challenges (Table 3). Codes that addressed potential problems associated with existing, popular LC supporting mechanisms are highlighted in bold, including “Refusal of or unsuitable adjustments”, “Unrealistic phased return”, and “Neglecting advice or guidance”.

**Table 3:** Workplace challenges for PwLC.

|  |  |  |
| --- | --- | --- |
| Phase 1<br>Returning or<br>preparing for<br>return | Absence hearings or procedures | P1, P3, P7-8, P13, P18 |
|  | Pressure to return to work (or return in person) | P3, P8, P13, P15 |
|  | **Unrealistic phased return** | **P3, P13-15, P17-18** |
|  | **Refusal of or unsuitable adjustments** | **P1-3, P12-15, P17-18** |
| Phase 2<br>When have<br>returned | Lack of recognition, awareness or understanding | P1-4, P6, P8, P11-16, P18-20 |
|  | Impact of work on Long COVID symptoms | P1, P3, P8, P10-19 |
|  | Excessive workload or responsibilities | P1-2, P10-11, P15-18 |
|  | **Neglecting advice or guidance** | **P1-3, P12-14, P18** |
|  | Discrimination or intimidation | P1, P13, P15, P18 |
| Workplace<br>Environment | Unsupportive culture | P2, P4, P12, P14-15, P18 |
|  | Existing policies and guidance | P2-4, P7-8, P11, P13-14, P17-18 |

##### Phase 1 Returning or preparing for return

Participants frequently reported engaging with the workplace to discuss RTW plans prior to resuming work. At this stage, challenges included absence hearings or procedures, deciding when to return, determining how to return, or, in cases of not returning, arranging sick leave and managing the impact of prolonged absence.

###### Absence hearings or procedure

Many participants had the experience of going through absence hearings, meetings or procedures. P1 told us that he had three hearings, and in the last hearing the Human Resources (HR) director explicitly stated they did not believe he had LC. The participant feared the procedure would eventually end his career. P18 reported anxiety associated with absence hearings, with her manager informing her that:

> *“…stage three [of the absence procedure]…is basically we are going to sack you”*

And that:

> *“…in reality 99% of people that go to stage three do lose their job.”*

In P7’s workplace, they simply called the procedure “*the get-rid-of-you process*”. Absence procedures themselves posed health effects on participants. Participants reported mental strain and physical demands that might worsen their LC symptoms. These included anticipated outcomes of absence procedures, the burden of managing unexpected situations, requirement to advocate for themselves, reviewing lengthy documents within tight deadlines, and confronting HR or senior management (P7, P8, P13).

###### Pressure to RTW (or Return in Person)

Many participants experienced pressure to RTW from the workplace, with pressure coming from line managers, HR, senior management, and sometimes colleagues or teams struggling with workload demands. Some participants were threatened with job loss or formal absence procedures, posing a major challenge for PwLC (P3, P13). Sometimes, the pressure was centred on returning in-person. For example, P1 felt he could have worked effectively from home with flexible schedules, but the company insisted on set hours in the office. He was also unable to cope with the daily commute.

###### Unrealistic phased return

Many participants reported shorter phased returns or quicker increases in working hours that were incompatible with their fluctuating or delayed recovery from LC (P3, P13-15, P17-18). For example, P13 described her experience with a proposed phased return:

> *“The doctor had said that I had to…that in her opinion she suggested at least two mornings a week to start with. The [senior officer], who’s not medically trained at all, said that wasn’t good enough and I had to start on…I think she said at least 20 hours a week to start with….They wanted me to try and build it up quite quickly, so they wanted me to do a phased return to go from, like, zero hours to 40 hours a week within the space of ten weeks and I just said, that’s a physical impossibility for me.”*

P15 said:

> *“I think the hardest things for HR and managers is the unpredictable nature of long COVID… But having a four week phased return is just not doable.…I think you’re setting people up to fail when you do that without a doubt”.*

One participant reported utilising their annual leave to obtain adequate rest (P18).

###### Refusal of or unsuitable adjustments

Participants reported workplace refusals to accommodate LC and make the adjustments they needed. Refusal included not supporting remote working (P1, P15) or flexible hours (e.g. taking breaks) (P1, P3), rejecting phased return plans suggested by Occupational Health or other healthcare professionals (P17), refusal to reduce job duties (P12, 18), requiring return to more physically demanding roles including night shifts (P13), or refusal to take a personalised approach that aligns work with an individual’s capacity or strengths (P2, P14). P15 recounted being required to drive a long distance on a non-working day for face-to-face training which was unrealistic for her, given her LC conditions. Similarly, P13 described the impact on her when asked to do early morning shifts:

> *“…they wanted me to start at 8 o’clock in the morning. So, to get there for 8 o’clock, to get up, to get showered…because to have a shower I have to use a bath aid to sit down in the bath, or in the shower, and then I have to sit down to have a rest, then get ready. I was having to get up at half past five to make sure I’ve got there. So, then they want me to do a morning’s work and then drive back home again – so, I’m physically exhausted, the fatigue is just off the scale.”*

In some cases, the manager agreed to the phased return plan and reduced hours but did not provide support or resources to share the original workload. P18 explained the situation:

> *“I have gone back, it is a full time job but I am doing four hours a week at the moment. He [the line manager] has not got anyone else to do any of the other parts of my job….He originally said that the first four weeks I would just be doing catching up… and doing my mandatory training. I wouldn’t have to have any contact with anyone….Then when I got back, he emailed everybody saying [participant’s name] is back now, get in touch with her about any issues… Technically I am doing three and a half day job in four hours.”*

##### Phase 2 When have returned

###### Lack of recognition, awareness or understanding

A lack of recognition, awareness or understanding of LC as a condition was the most commonly reported workplace challenge among participants. Fifteen participants shared such experiences (Table 3). The most significant problem participants reported was scepticism regarding LC as a recognised condition (P1-2, P6, P13). Participants described the “*If you can’t see it, it doesn’t exist”* (P2) attitude from management and colleagues as most of the symptoms and debilitating effects of LC were hidden.

> *“…They’ll be like, oh you’re looking really well, when are you coming back to do x, y and z. And it would have been easier if I’d had a limb in a cast or something like that, or some scar I could show them.”* (P8)

Sometimes it was because people could not tell the difference between LC symptoms and general malaise, such as differentiating fatigue from tiredness (P14, P18-19). Another challenge to achieving a good understanding of LC was the expected rate of recovery versus potential symptom relapse combined with the sometimes slow, unpredictable nature of LC (P2-3, P6, P8, P12-13, P16, P18, P20). For example,

> *“I don’t think my managers can get their head round the fact that I’m not that person anymore. They seem to think that I’ll be okay in a month or it’s just a phase… They’re just not listening.”* (P2)

> *“…I keep getting called out by my line manager saying I’m not working hard enough and it’s not fair on the rest of the team that I get to go away from my desk for five minutes every couple of hours…”* (P12)

###### Impact of work on LC symptoms

The impact of work on LC was another major challenge highlighted by participants. When felt pushed to work, many experienced deterioration in physical, cognitive, emotional, and psychological symptoms. Participants reported prolonged recovery time associated with these symptoms impacting their personal or home life. For example:

> *“I thought, okay, I must be better. I did a week of six small training sessions …and I just really crashed and I had really bad PEM (post-exertional malaise), felt terrible. So, that’s like really bad fatigue, really bad nausea, really bad brain fog, you feel like your whole immune system is going crazy… I’m really sick.”* (P16)

> *“… because I work, I’m finding that on my days off, at weekend, I’m spending most of my time in bed, and then get resentful, because I feel like I’m giving everything to work and there’s nothing left for me….weekends…I haven’t got the energy to do anything.”* (P2)

> *“It’s my cognitive function, it became clear the harder I tried to work and just push through, the worse I got. I just got worse and worse basically.”* (P15)

> *“The crash that I had,… like, when I had mild symptoms at the start, the thing which really wrecked me was doing a day of work where I didn’t move, I just spoke to people and tried to use my brain and it completely destroyed me.”* (P19)

###### Excessive workload or responsibilities

Participants reported challenges to RTW related to excessive workload or responsibilities, and employers being unwilling or unable to make adjustments. Some faced high workloads because the employer refused to adjust (P1), was unable to make adjustments despite wanting to do so (P15), or were not able to provide adequate resources (P18). Participants attributed this to stressful or competitive work culture (P11, P16), or a rigid organisational culture that struggled to adapt to individual health needs (P2, P17). For some participants their LC symptoms impacted their ability to perform safety or time critical job roles, for example participants found the physical or cognitive load required to respond to emergencies and make rapid decisions particularly challenging (P10, P15).

###### Neglecting advice or guidance

Participants reported advice and guidance from occupational health or other health professionals was not always followed. Participants felt pressured to follow the organisation or company’s requests (P1, P14), returning to work when they were still assessed as unfit for work (P13), working more hours than advised (P13), rushing phased return plans (P3, P12), or refusal to offer flexibility (P18). For example,

> *“Occupational health had said, no, you’re not fit to return to work. The long COVID doctor had said, you’re not fit to return to work. But work was still hounding me to say, we need you to come back to work.”* (P13)

> *“There was the plan that if I struggled at any week with that number of hours, then we’d go back a week and go down in hours and stuff, but that didn’t get followed. And so, then I was rushed through that phased return and that’s when everything started going downhill.”* (P12)

###### Discrimination or intimidation

Some participants reported experiencing discrimination because of their LC condition (P1, P15, P18). In rare cases, the participants faced intimidation such as being shouted at during absence meetings (P1), supportive colleagues being told by senior management to “*stay out of it*” (P1), being told they would be managed out of the job (P18) or participants being warned to remain silent to avoid conflict with senior management (P13).

##### Workplace environment

###### Unsupportive culture

Several participants described the work environment as unsupportive, creating strained working relations with managers and among colleagues, and frustration in communication. For example,

> *“I’ve tried everything, I just don’t know what else to do, and you’d think the government… I don’t know whether it’s local management or my employer as a whole, I don’t know.”* (P2)

> *“So, most of the people don’t get it at all and are quite happy to tell you that they think you’re faking.”* (P12)

> *“I think there is a lot of burnout and a lot of people working beyond.… I think four years ago would have been a lot more supportive…they are, like, their own situation of being exhausted, fatigued, but just, sort of, burnt out basically… I think that is a cultural shift.”* (P18)

###### *Existing* policies and guidance

Several participants found that existing policies were inflexible, did not work for LC, or was even used against them (P2-4, P7-8, P11, P13-14, P17-18). Examples included national tax rules (P7), pension rules (e.g. medical retirement) (P4), organisation-wide policy on sick leave and phased return (P2-3, P8, P13-14, P17). P18 expressed concern over a capability policy in her organisation that could be used to terminate her contract. P14 suggested that the organisational policy was too inflexible and must be reformed to better support people working with long-term conditions, such as LC, mental health issues and other chronic conditions. P7, who was on a fixed-term contract, reported that the company stopped renewing it because “*they had to have somebody who was well in that job*”.

###### Government support

The UK government support, such as Personal Independence Payments in England (PIP, or Adult Disability Payment in Scotland), Access to Work, and Employment and Support Allowance (ESA), played an important role in supporting PwLC to RTW.

However, participants talked about the emotional burden and stress of navigating the application process. P15 described it as such:

> *“…it’s degrading, it’s mentally exhausting. And there are so many hoops to jump through to actually get those benefits.”*

P7 said:

> *“…the completing of the forms is painful and it’s very lengthy and very difficult, because you don’t know what they want of you, and none of it makes sense. And it’s in a very negative experience because all you’re doing is focusing on what’s wrong with you.”*

As a result, not all participants succeeded in getting government support.

> *“I keep on trying to apply for PIP and everything but DWP don’t seem very impressed. And so…I’ve got another tribunal coming up, which is fairly traumatic… They just like catching you out. They have all sorts of traps to set for me, so it’s been a struggle and I just don’t think I’ll get any help anywhere.”* (P1)

Department for Work and Pensions or DWP is the UK government department responsible for work-related benefits and support. Those who received benefits expressed appreciation for the support provided. Participants also appreciated the help the LC Clinic and Citizens Advice provided for them in getting the government support (P9, P13, P15).

### The Managers’ Perspective

#### Manager Perspective - Support

Both participants demonstrated strong support for employees with LC, doing everything possible to help them return to work and remain in their roles. However, their approaches differed due to the different sizes of their organisations. M1 described a comprehensive supporting system where guidance, procedures, risk assessment, work adjustments, referral for treatment and rehabilitation were all in place. Conversely, M2 took a more hands-on role, overseeing the whole process and was more able to negotiate directly with the organisation to gain “bespoke support” for the employee, e.g. paid leave for therapies.

#### Manager perspective - Challenges

M1 discussed several challenges while supporting employees with LC. One key issue was securing approval for extended phased return. While she recognised that some employees with LC needed longer than the standard four-week phased return, final approval rested with others in the organisation. Additionally, work adjustments were not always possible where roles could not easily be fulfilled through remote work or desk work. M1 also admitted having limited follow-up capacity due to high case load, which hindered opportunities for her to learn and improve support strategies.

Workplace relationship challenges, stemming from perceived preferential treatment, added to the complexity. Another challenge involved delays in access to specialist medical care and subsequent advice impacting on appropriate RTW support.

M2 found the lack of formal policy and specific guidance for accommodating employees with LC a major challenge. Additionally, the variability in symptoms presented another challenge:

> *“…it displays in many different ways…even though there’s some commonalities, and I think because COVID changed and changed and changed, …one person’s symptoms is not going to be the same as the next, so you have to greet each person with an open mind to…to sort of understand their difficulties.”* (M2)

M2 reported that stigma associated with LC prevented one former employee from seeking support, eventually opting for early retirement. In light of the absence of a supporting structure for PwLC, M2 explained that she was able to arrange paid leave for an affected employee, only due to the small size of the organisation and personal connections, coupled with senior management’s favourable view of the employee’s past contributions. M2 also stated that financial constraints, insufficient resources, job demand, and lack of options for remote working presented challenges for her to effectively support employees with LC to RTW. These challenges corresponded with concerns raised by PwLC participants.

Both managers mentioned the importance of being creative with existing guidance to develop personalised approaches for accommodating employees with LC.

> *“Yes [follow the guidance], and be creative to look at [individual needs]… So we’d work alongside there. They could say things like they need staff car parking close to the building, they need something a bit more bespoke than just the generic sort of practical advice.”* (M1)

> *“I think other than a doctor’s recognising that Long COVID was possibly an issue, it [effective support] didn’t really come from, I don’t believe, a medical opinion. It came from the school and her working together to work out how we were going to get her back to work.”* (M2)

## Discussion

This qualitative research explored both perspectives of PwLC and managers to understand what support has been provided to PwLC in the UK in relation to RTW, and challenges and barriers associated with existing support mechanisms.

### A personalised approach for supporting PwLC to RTW

This study highlights the specific difficulties faced in managing RTW with LC. LC symptoms are diverse and can fluctuate, creating specific challenges for the supporting systems. A tailored approach, including flexible schedules, remote work options, and restructured job roles to align with physical and cognitive capacities, were key facilitators for PwLC to RTW. Previous research has reported challenges with RTW programmes being too rigid to accommodate the fluctuating symptoms of LC, complicating the balance between work and health recovery [1, 13]. When extended phased returns recommended by Occupational Health were not implemented, or workplace adjustments were insufficient or unsuitable for the individual, workers faced significant challenges, including exhaustion, difficulty maintaining a work-life balance, and, in some cases, relapses of their condition [14, 25, 26]. During discussions about the shortcomings of existing policies and guidance, both PwLC and managers highlighted the necessity of a personalized approach. This perspective aligns with existing evidence, which emphasises the importance of RTW strategies tailored to the individual’s specific symptoms and recovery needs, acknowledging the variable and unpredictable nature of LC [16, 26].

Phased returns, reduced hours, occupational health services, and job adjustments are existing support mechanisms for RTW with long-term conditions, but our study highlights the challenges caused by impersonal or inconsistent application. Phased return plans, for instance, were found to be unrealistic in some cases, with expectations for recovery timelines that failed to account for the unpredictable and fluctuating nature of LC. Although Occupational Health services played a critical role in providing guidance to employers, their recommendations were not always implemented effectively. Work adjustments, if not aligned with individual’s pace or stage of recovery, could fail to provide adequate support. This mirrors previous study findings suggesting that flexibility is important to sustain RTW for PwLC, and that rehabilitation programmes focussing on health alone may not be sufficient [27].

### Education, training and guidance

The need for education and training was emphasised by both PwLC and managers. As a new condition, LC remains poorly understood in terms of its underlying pathophysiology and symptoms, as noted in NICE guidance [28]. Participants highlighted significant knowledge and guidance gaps at individual and organisational levels: the lack of knowledge related to LC itself, managing LC in the context of RTW, and RTW policies more generally. Managers reported that the lack of guidance and information about LC left them feeling ill-equipped to address the unique needs of affected employees, echoing the findings of Bramwell et al [19]. Several national guidance on LC have become available in the UK since 2021, e.g. the Faculty of Occupational Medicine (FOM) guidance on RTW in LC [29], the Society of Occupation Medicine (SOM) guidance [30], and that from the Chartered Institute of Professional Development (CIPD) [31]. To ensure the guidance are understood and followed, education and training must be provided to organisations and managers who are managing PwLC on daily basis. This aligns with previous research, which describes the importance of enhancing awareness about LC among all actors involved in the RTW process, as well as improving communication between them to support effective reintegration [1, 9, 13, 26].

Several participants reported concerns about stigma associated with a LC diagnosis and subsequent negative impact on RTW. This is likely resulted from a stressful work culture and the lack of awareness and understanding about LC symptoms in workplaces, findings which have been replicated elsewhere [16, 17]. As most of the LC symptoms are not as visible as other physical disabilities, participants found the needs to advocate for themselves in order to gain better understanding. It highlights the importance of a supportive environment where PwLC would not hesitate to communicate openly about their conditions and seek support. This echoes what has been found by Nielsen and Yarker [8] that workers with LC who received understanding and support from their colleagues and managers were better able to navigate their RTW journey.

### The role of government support

Both PwLC and managers experienced limited resources and inflexible policies hindering the development of personalised solutions. For PwLC, financial pressure is a main personal factor to RTW, which increases the risk of presenteeism and exacerbation of symptoms. In the UK, financial support for PwLC is primarily through statutory benefits. However, participants talked about the physical and emotional burden of navigating the welfare systems and application process. For managers, some of the adjustments they make, e.g. reducing working hours, or change of job roles could have financial consequences for PwLC. Furthermore, providing tailored support for PwLC entails mobilisation of resources within the organisation, e.g. extending phased return or arranging backup personnel. Therefore, there is a role for the government to play to fill the gap and provide additional resources to support individuals and employers.

The UK government has recently announced significant changes in welfare system that aim to facilitate people with long-term conditions to RTW and hence reduce the benefit budget [32]. It is stated that the reform will reduce the complexity in obtaining support for those have severe disabilities, ensure the ‘right to try’ to work without losing benefits, and invest substantially in Job Centres and work coaches to develop tailored support. Furthermore, there have been changes to sick pay that removed statutory sick pay rebate for Small and Medium Enterprises (SMEs) which may have negatively impacted disproportionally PwLC who work for SMEs [33]. As each individual’s experience with LC is unique, with varying symptoms, recovery trajectories, and personal circumstances, the welfare system reform should take this into account to ensure equitable support for PwLC and other long-term conditions to RTW.

### Strengths and limitations

A key strength of our study is a comprehensive exploration of the PwLC perspective on supporting RTW. The addition of the managers’ perspective was also beneficial, however given the time constraint we only interviewed two managers with such experiences. A PPI representative has been advising the research team throughout the process, which is another strength of the study. The study is timely as the UK government is reforming its welfare system to better support people with long-term conditions to RTW.

We achieved a good coverage in terms of company size and job sectors. However, there is a lack of diversity in terms of education levels and ethnicity. Most of the participants held a university degree and only one participant was considered to belong to an ethnic minority group. Some of our recommendations highlight the role of Occupational Health. Nevertheless, we are aware that access to occupational health services is not universal in the UK and hence many workers, particularly those working in SMEs may not have access to this support.

## Conclusion

This study highlights the need for personalised and flexible workplace accommodations to support PwLC, addressing challenges like fluctuating symptoms and financial constraints. Education and training for managers are vital to improving understanding and implementation of support policies. Encouraging changes at an organisational and policy level, will create a more inclusive and effective work environment for PwLC.

## Declarations

### Ethics approval and consent to participate

Ethical approval was obtained from the University of Manchester Research Ethics Committee (Ref: 2024-17658-32518). All participants received Participant Information Sheet at one working day before the interviews and provided their consent to participate.

### Consent for publication

All participants consented for publication of anonymised results.

### Availability of data and materials

Data is provided within the manuscript or supplementary information files.

### Competing interests

None declared.

### Funding

This research was funded by the University of Manchester Research Institute (UMRI).

### Authors’ contributions

HW and SD conducted the interviews. HW, SD and RW drafted the manuscript. MvT, AC, DB, and DM contributed to study design, interview schedule development and data analysis. DF provided advises to the research as a patient and public involvement (PPI) representative. All members of the research team reviewed and commented on each version of the manuscript.

## Acknowledgements

We thank members of the LC Patient Support Group Greater Manchester for their advice to the study design, participant engagement and recruitment.

## Notes

### Competing Interest Statement

The authors have declared no competing interest.

### Author Declarations

Ethical approval was granted by the University of Manchester Research Ethics Committee (Ref: 2024-17658-32518). All participants received Participant Information Sheet at one working day before the interviews and provided their consent to participate.

## Reference

1. Lunt J, Hemming S, Elander J, et al. Sustaining work ability amongst female professional workers with long COVID. Occupational Medicine. 2024;74(1):104–12.

2. Chasco EE, Dukes K, Jones D, et al. Brain Fog and Fatigue following COVID-19 Infection: An Exploratory Study of Patient Experiences of Long COVID. International Journal of Environmental Research and Public Health. 2022;19(23):15499.

3. Gyllensten K, Holm A, Sandén H. Workplace factors that promote and hinder work ability and return to work among individuals with long-term effects of COVID-19: A qualitative study. Work. 2023;75:1101–12.

4. Davis HE, McCorkell L, Vogel JM, et al. Long COVID: major findings, mechanisms and recommendations. Nature Reviews Microbiology. 2023;21(3):133–46.

5. Office for National Statistics. Self-reported coronavirus (COVID-19) infections and associated symptoms, England and Scotland: November 2023 to March 2024 2024 [Available from: https://www.ons.gov.uk/peoplepopulationandcommunity/healthandsocialcare/conditionsanddiseases/articles/selfreportedcoronaviruscovid19infectionsandassociatedsymptomsenglandandscotland/november2023tomarch2024.

6. Woodrow M, Ziauddeen N, Smith D, et al. Exploring Long Covid Prevalence and Patient Uncertainty by Sociodemographic Characteristics Using GP Patient Survey Data. Health Expectations. 2025;28(2):e70202.

7. Saade A, Didier Q, Cha L, et al. The prevalence, determinants, and consequences of post-COVID in healthcare workers: A cross-sectional survey. Journal of Medical Virology. 2024;96(6):e29725.

8. Nielsen K, Yarker J. “It’s a rollercoaster”: the recovery and return to work experiences of workers with long COVID. Work & Stress. 2024;38(2):202–30.

9. McNabb KC, Bergman AJ, Smith-Wright R, et al. “It was almost like it’s set up for people to fail” A qualitative analysis of experiences and unmet supportive needs of people with Long COVID. BMC Public Health. 2023;23(1):2131.

10. Folk AL. Exploring the experiences of academic library employees with long COVID in the United States and Canada. The Journal of Academic Librarianship. 2023;49(6):102790.

11. Pauwels S, Boets I, Polli A, et al. Return to work after long COVID: evidence at 8th March 2021. Health and Safety Executive; 2021. Contract No.: ER003 (2021) Evidence Report.

12. Daniels S, Wei H, McElvenny DM, et al. Return to Work with Long COVID: A Rapid Review of Support and Challenges. medRxiv. 2025:2025.04.29.25326647.

13. Kohn L, Dauvrin M, Detollenaere J, et al. Long COVID and return to work: a qualitative study. Occup Med (Lond). 2024;74(1):29–36.

14. Delgado-Alonso C, Cuevas C, Oliver-Mas S, et al. Fatigue and Cognitive Dysfunction Are Associated with Occupational Status in Post-COVID Syndrome. International Journal of Environmental Research and Public Health [Internet]. 2022; 19(20).

15. Schmachtenberg T, Müller F, Kranz J, et al. How do long COVID patients perceive their current life situation and occupational perspective? Results of a qualitative interview study in Germany. Front Public Health. 2023;11.

16. Miller A, Song N, Sivan M, et al. Identifying the needs of people with long COVID: a qualitative study in the UK. BMJ Open. 2024;14(6):e082728.

17. Damant RW, Rourke L, Cui Y, et al. Reliability and validity of the post COVID-19 condition stigma questionnaire: a prospective cohort study. eClinicalMedicine. 2023;55.

18. Gamillscheg P, Łaszewska A, Kirchner S, et al. Barriers and facilitators of healthcare access for long COVID-19 patients in a universal healthcare system: qualitative evidence from Austria. International Journal for Equity in Health. 2024;23(1):220.

19. Bramwell D, Sanders C, Rogers A. A case of tightrope walking: An exploration of the role of employers and managers in supporting people with long-term conditions in the workplace. International journal of workplace health management. 2016;9(2):238–50.

20. Tong A, Sainsbury P, Craig J. Consolidated criteria for reporting qualitative research (COREQ): a 32-item checklist for interviews and focus groups. International Journal for Quality in Health Care. 2007;19(6):349–57.

21. Tolley EE, Ulin PR, Mack N, et al. Qualitative methods in public health: a field guide for applied research: John Wiley & Sons; 2016.

22. Fleiss JL, Levin B, Paik MC. Statistical methods for rates and proportions: john wiley & sons; 2013.

23. Braun V, Clarke V. Reflecting on reflexive thematic analysis. Qualitative research in sport, exercise and health. 2019;11(4):589–97.

24. Braun V, Clarke V. Using thematic analysis in psychology. Qualitative research in psychology. 2006;3(2):77–101.

25. Schmid S, Uecker C, Fröhlich A, et al. Effects of an integrative multimodal inpatient program on fatigue and work ability in patients with Post-COVID Syndrome—a prospective observational study. European Archives of Psychiatry and Clinical Neuroscience. 2024.

26. Straßburger C, Hieber D, Karthan M, et al. Return to work after Post-COVID: describing affected employees’ perceptions of personal resources, organizational offerings and care pathways. Front Public Health. 2023;11.

27. Brehon K, Niemeläinen R, Hall M, et al. Return-to-Work Following Occupational Rehabilitation for Long COVID: Descriptive Cohort Study. JMIR Rehabil Assist Technol. 2022;9(3):e39883.

28. National Institute for Health and Excellence. Long-term effects of coronavirus (long COVID): What is it? 2022 [Available from: https://cks.nice.org.uk/topics/long-term-effects-of-coronavirus-long-covid/background-information/definition/.

29. FOM. Guidance for managers and employers on facilitating return to work of employees with long-COVID. In: Medicine TFoO, editor. Online: The Faculty of Occupational Medicine; 2021.

30. The Society of Occupational Medicine. Long COVID and Return to Work – What Works?2022 13 May 2024. Available from: https://www.som.org.uk/sites/som.org.uk/files/Long_COVID_and_Return_to_Work_What_Works.pdf.

31. CIPD. Working with long COVID: Research evidence to inform support. London: Chartered Institute of Personnel and Development.; 2022.

32. DWP. Spring Statement 2025 health and disability benefit reforms - Impacts In: Pensions DfWa, editor. London: DWP.

33. Mayne M. What does the NHS long Covid pay decision mean for employers? People Management [Internet]. 2023. Available from: https://www.peoplemanagement.co.uk/article/1812127/does-nhs-long-covid-pay-decision-mean-employers.

