## Supplementary File 1 and 2 for "Long COVID and Work in the UK: Challenges, Support and Perspectives"

### Supplementary file 1 interview questions

**Proposed questions for UMRI Long COVID interviews – people with Long COVID**

**A. Questions for People with Long COVID**

Brief medical background:

- Can you tell us briefly when and how your Long Covid was diagnosed?
- What are the main symptoms? Any associated conditions eg POTS (Postural tachycardia syndrome)/chronic fatigue? (if the interviewee asks about the terms, we can explain)
- Do you have other long-term conditions?

General work history

- Which sector or industry do you normally work in?
- Can you tell us about your pre-COVID employment? (employer, relationship with employer / line manager, size of company, role and duties, work flexibility (pre and post pandemic), contract type)
- Capacity to work at home/flexibility/furloughed / redundancy
- Time off work with Covid infection/Long COVID
- Knowledge around sickness policy and entitlement to sick pay.

Post COVID employment and relevant changes

- Have you returned to work or not, or returned with a different arrangement? If returned differently, what are the changes? (job, tasks, patterns/working hours)
- Are these changes mainly or partly due to Long Covid? How were you supported to RtW (if this occurred)
- How does current situation (either in work or not) match your expectations? If not quite, is it due to your Long Covid symptoms or some other reason?

Suggestions for improvement

- Have you taken time off work as result of Long COVID? If yes, barriers to returning to work and facilitators for helping return to work.
- If no time taken off work, what supported your ability to stay in work? (support, adaptations etc)?
- Support from employer – who from, how was this structured, any changes to relationships at work (employer, line manager, colleagues).
- What support would you have liked but didn’t get?

What support received other than via employment?

- Support from medical staff (GP, hospital, Long COVID clinic etc)
- Support from family, friends, other agencies
- Support from patient support groups, community groups, charities, etc
- Union involvement
- Benefits/financial support

General

- Awareness of any specific guidance for helping with return to work with/after Long COVID (national, regional, company) – engaged with your employer about this, helpfulness.
- If aware, do you think any of the guidance are helpful? If yes can you specify?

**Proposed questions for UMRI Long COVID interviews – Employers/ Human Resources / line managers**

**B. Questions for Employers/HR/managers**

Understand barriers/facilitators to return to work from a human resources/line management perspective, exploring use of existing return to work guidance for Long COVID

- Size of organisation, sector, industry
- Any personal experiences of Long COVID – self /family /friends /employees? And/or understanding of Long COVID as a condition
- Awareness of any specific guidance for helping with Return to work after / with Long COVID (national, regional)
- What are the company policies and practices related to Long COVID [ask for a copy], including vaccination
- What are the company policies and practices related to health and wellbeing [ask for a copy]
- Sickness absence and pay for Long COVID. Has this changed over the period, and has it influenced return to work practices in your organization?
- Any informal practices at the workplace for support e.g. staff groups, Unions, local community
- Experience of supporting those with Long COVID return to work
  - What do you see as your role in supporting people with Long COVID to return to and remain in work?
  - If supported more than one employee, how has your experience in supporting them differed and why?
- Barriers and facilitators for helping people after / with Long COVID return to work
- Have different groups of workers experienced the return to work after / with Long COVID differently (full time vs part time, caring responsibilities, age, gender, ethnicity, other).
- Suggestions for improving the Return to work process (in relation to Long COVID) – what support would/do you need as employer /human resources/ line manager?

### Supplementary file 2: Consolidated criteria for reporting qualitative studies (COREQ): 32-item checklist

Developed from:

Tong A, Sainsbury P, Craig J. Consolidated criteria for reporting qualitative research (COREQ): a 32-item checklist for interviews and focus groups. *International Journal for Quality in Health Care*. 2007. Volume 19, Number 6: pp. 349 – 357

| **No. Item** | **Guide questions/description** | **Comments** |
| --- | --- | --- |
| **Domain 1: Research team and reﬂexivity** | | |
| *Personal Characteristics* | | |
| 1. Interviewer/facilitator | Which author/s conducted the interview or focus group? | In “Recruitment and Data Collection” |
| 2. Credentials | What were the researcher’s credentials? E.g. PhD, MD | In “Recruitment and Data Collection” |
| 3. Occupation | What was their occupation at the time of the study? | In “Recruitment and Data Collection” |
| 4. Gender | Was the researcher male or female? | HW and SD are females |
| 5. Experience and training | What experience or training did the researcher have? | In “Recruitment and Data Collection” |
| *Relationship with participants* | | |
| 6. Relationship established | Was a relationship established prior to study commencement? | No |
| 7. Participant knowledge of the interviewer | What did the participants know about the researcher? e.g. personal goals, reasons for doing the research | In “Recruitment and Data Collection” |
| 8. Interviewer characteristics | What characteristics were reported about the interviewer/facilitator? e.g. Bias, assumptions, reasons and interests in the research topic | Research objective stated in “Introduction” |
| **Domain 2: study design** | | |
| *Theoretical framework* | | |
| 9. Methodological orientation and Theory | What methodological orientation was stated to underpin the study? e.g. grounded theory, discourse analysis, ethnography, phenomenology, content analysis | In “Data analysis” |
| *Participant selection* | | |
| 10. Sampling | How were participants selected? e.g. purposive, convenience, consecutive, snowball | In “Recruitment and Data Collection” |
| 11. Method of approach | How were participants approached? e.g. face-to-face, telephone, mail, email | In “Recruitment and Data Collection” |
| 12. Sample size | How many participants were in the study? | In “Methods” |
| 13. Non-participation | How many people refused to participate or dropped out? Reasons? | In “Recruitment and Data Collection” |
| *Setting* | | |
| 14. Setting of data collection | Where was the data collected? e.g. home, clinic, workplace | In “Recruitment and Data Collection” |
| 15. Presence of non-participants | Was anyone else present besides the participants and researchers? | No |
| 16. Description of sample | What are the important characteristics of the sample? e.g. demographic data, date | In “Demographics” |
| *Data collection* |  |  |
| 17. Interview guide | Were questions, prompts, guides provided by the authors? Was it pilot tested? | In Supplementary File 1 |
| 18. Repeat interviews | Were repeat interviews carried out? If yes, how many? | No |
| 19. Audio/visual recording | Did the research use audio or visual recording to collect the data? | In “Recruitment and Data Collection” |
| 20. Field notes | Were ﬁeld notes made during and/or after the interview or focus group? | Yes |
| 21. Duration | What was the duration of the interviews or focus group? | In “Recruitment and Data Collection” |
| 22. Data saturation | Was data saturation discussed? | In “Recruitment and Data Collection” |
| 23. Transcripts returned | Were transcripts returned to participants for comment and/or correction? | No |
| **Domain 3: analysis and ﬁndings** | | |
| *Data analysis* | | |
| 24. Number of data coders | How many data coders coded the data? | Two |
| 25. Description of the coding tree | Did authors provide a description of the coding tree? | In “Results” |
| 26. Derivation of themes | Were themes identiﬁed in advance or derived from the data? | In “Data analysis” |
| 27. Software | What software, if applicable, was used to manage the data? | In “Data analysis” |
| 28. Participant checking | Did participants provide feedback on the ﬁndings? | No |
| *Reporting* | | |
| 29. Quotations presented | Were participant quotations presented to illustrate the themes/ﬁndings? Was each quotation identiﬁed? e.g. participant number | Yes and Yes. In “Results” |
| 30. Data and ﬁndings consistent | Was there consistency between the data presented and the ﬁndings? | Yes. |
| 31. Clarity of major themes | Were major themes clearly presented in the ﬁndings? | Yes. In “Results” |
| 32. Clarity of minor themes | Is there a description of diverse cases or discussion of minor themes? | Yes. In “Results” |
